# Risk Factors Associated with Clinical Outcomes in 323 COVID-19 Patients in Wuhan, China

**DOI:** 10.1101/2020.03.25.20037721

**Authors:** Ling Hu, Shaoqiu Chen, Yuanyuan Fu, Zitong Gao, Hui Long, Hong-wei Ren, Yi Zuo, Huan Li, Jie Wang, Qing-bang Xu, Wen-xiong Yu, Jia Liu, Chen Shao, Jun-jie Hao, Chuan-zhen Wang, Yao Ma, Zhanwei Wang, Richard Yanagihara, Jian-ming Wang, Youping Deng

## Abstract

**Background:** With evidence of sustained transmission in more than 190 countries, coronavirus disease 2019 (COVID-19) has been declared a global pandemic. As such, data are urgently needed about risk factors associated with clinical outcomes.

**Methods:** A retrospective chart review of 323 hospitalized patients with COVID-19 in Wuhan was conducted. Patients were classified into three disease severity groups (non-severe, severe, and critical), based on their initial clinical presentation. Clinical outcomes were designated as favorable and unfavorable, based on disease progression and response to treatments. Logistic regression models were performed to identify factors associated with clinical outcomes, and log-rank test was conducted for the association with clinical progression.

**Results:** Current standard treatments did not show significant improvement on patient outcomes in the study. By univariate logistic regression model, 27 risk factors were significantly associated with clinical outcomes. Further, multivariate regression indicated that age over 65 years, smoking, critical disease status, diabetes, high hypersensitive troponin I (>0.04 pg/mL), leukocytosis (>10 × 10^9^/L) and neutrophilia (>75 × 10^9^/L) predicted unfavorable clinical outcomes. By contrast, the use of hypnotics was significantly associated with favorable outcomes. Survival analysis also confirmed that patients receiving hypnotics had significantly better survival.

**Conclusions:** To our knowledge, this is the first indication that hypnotics could be an effective ancillary treatment for COVID-19. We also found that novel risk factors, such as higher hypersensitive troponin I, predicted poor clinical outcomes. Overall, our study provides useful data to guide early clinical decision making to reduce mortality and improve clinical outcomes of COVID-19.

(Funded by the Natural Science Foundation of Hubei Province ZRMS2019000029 and the Top Youth Talent Program in Hubei Province.)

## INTRODUCTION

Coronavirus disease 2019 (COVID-19) is a potentially lethal respiratory illness caused by a newly identified coronavirus, named SARS coronavirus 2 (SARS-CoV-2), which was first recognized in December 2019 in Wuhan, in Hubei Province, China.^1,2^ The disease has spread rapidly to more than 190 countries, and as of March 24, 2020, 422,915 confirmed cases and 18,915 deaths have been officially reported worldwide.^3-7^ With sustained transmission on six continents, the World Health Organization (WHO) has recently declared COVID-19 as a global pandemic.

Most previous studies of COVID-19 have focused primarily on epidemiological and clinical characteristics.^8-12^ Wang and co-workers compared the clinical features of 138 hospitalized patients with non-severe and severe COVID-19.^10^ Guan and colleagues updated the clinical characteristic and disease severity in 1,099 laboratory-confirmed cases throughout China.^12^ Only a few studies have investigated risk factors and clinical outcomes.^13,14^ So, it is urgent to identify potential novel risk factors and treatments associated with patient-centered outcomes of COVID-19.

In this study, we analyzed the clinical course of 323 hospitalized patients with COVID-19 from January 8 to March 10, 2020, in Wuhan, to identify risk factors associated with clinical outcomes for improving management guidelines.

## METHODS

### Study design

The institutional ethics board of Tianyou Hospital, an affiliate of the Wuhan University of Science and Technology, approved the conduct of this retrospective review. Oral consent was obtained from patients and written informed consent was waived. Tianyou Hospital is one of several designated hospitals for the treatment of COVID-19 in Wuhan.

### Case definition

Electronic medical records (EMR) from inpatients with COVID-19 at Tianyou Hospital were studied. Diagnosis complied with the WHO interim guidance^15^ and the guidelines of COVID-19 diagnosis and treatment trial 5^th^ edition, by the National Health Commission of the People’s Republic of China.^16^

The COVID-19 diagnosis was based on (1) the exclusion of other known infectious and non-infectious causes of pneumonia; (2) exposure history in Wuhan in the most recent 14 days or contact history with a confirmed patient or disease cluster; and (3) clinical presentation of fever, respiratory symptoms, characteristic chest computer tomography (CT) image and/or leukopenia and lymphopenia.

### Data sources

A total of 323 patients were enrolled from January 8 to February 20, 2020. Real-time reverse transcription polymerase chain reaction (rRT-PCR) was performed on throat swab specimens of all patients. Clinical signs, disease onset, laboratory tests (including rRT-PCR and CT), treatments, co-morbidities, complications, and outcome data were collected from EMR. All raw data were initially assessed by trained physicians. Based on the clinical presentation at the time of admission, patients were categorized into one of three groups: non-severe, severe and critical. Clinical outcomes (favorable or unfavorable) were based on an average observation period of 28 days, with March 10, 2020 as the final follow-up date.

### Clinical classification

The disease severity groups included the following: non-severe (patients showed fever, respiratory symptoms and CT presentation of pneumonia; severe (patients showed respiratory distress with RR≥30 breaths/min, oxygen saturation less than 93%, arterial partial pressure of oxygen/oxygen concentration less than 300 mmHg); critical (patients showed respiratory failure requiring ventilatory support, as well as shock and organ dysfunction requiring intensive care).

### Clinical outcomes

Disease improvement or favorable clinical outcome included full recovery and discharge, progression from critical/severe to non-severe disease status, PCR positive to negative, and/or maintenance of non-severe status. Disease progression or unfavorable clinical outcome included death, progression from non-severe to severe/critical disease status or severe to critical status, and/or maintenance of severe or critical status.

### Statistics Analysis

Chi-square test or Fisher exact test was used for categorical variables measurements. For continuous variables, student T-test or Mann-Whitney test was used. Multiple imputation was conducted to handle missing data.^17^ Odds ratios (ORs) and 95% confidence intervals (CIs) were calculated using univariate and multivariate logistic regression models. For survival analysis, the survival time was defined as the interval from the date of admission to the date of death or discharge. The association of risk factors with clinical outcome was analyzed using the Kaplan-Meier method and log-rank test. All analyses were implemented with R software (version 3.6.2) or Statistical Analysis System (SAS) software (version 9.4, SAS Institute Inc., Cary, NC). All *P* values were two-sided, and those < 0.05 were considered as statistically significant.

## RESULTS

### Clinical characteristics

Of the 323 patients with COVID-19, 186 (57.6%) were rRT-PCR positive and 137 (42.4%) patients were rRT-PCR negative but had typical chest CT image, respiratory symptoms and compatible blood test results at the time of admission. At the end of the study, 252 patients had recovered and were discharged, 35 patients had died (overall case fatality rate, 10.8%), and 36 patients were still hospitalized.

Based on their initial clinical presentation, the 323 patients were classified into the non-severe (151), severe (146) and critical (26) disease groups (Table 1). There was no gender difference between the three groups. The median age of patients was 61 years (range, 23–91). Patients over 65 years were overrepresented within the severe (43.2%, 63/146) and critical (57.7%, 15/26) disease groups.

**Table 1.**
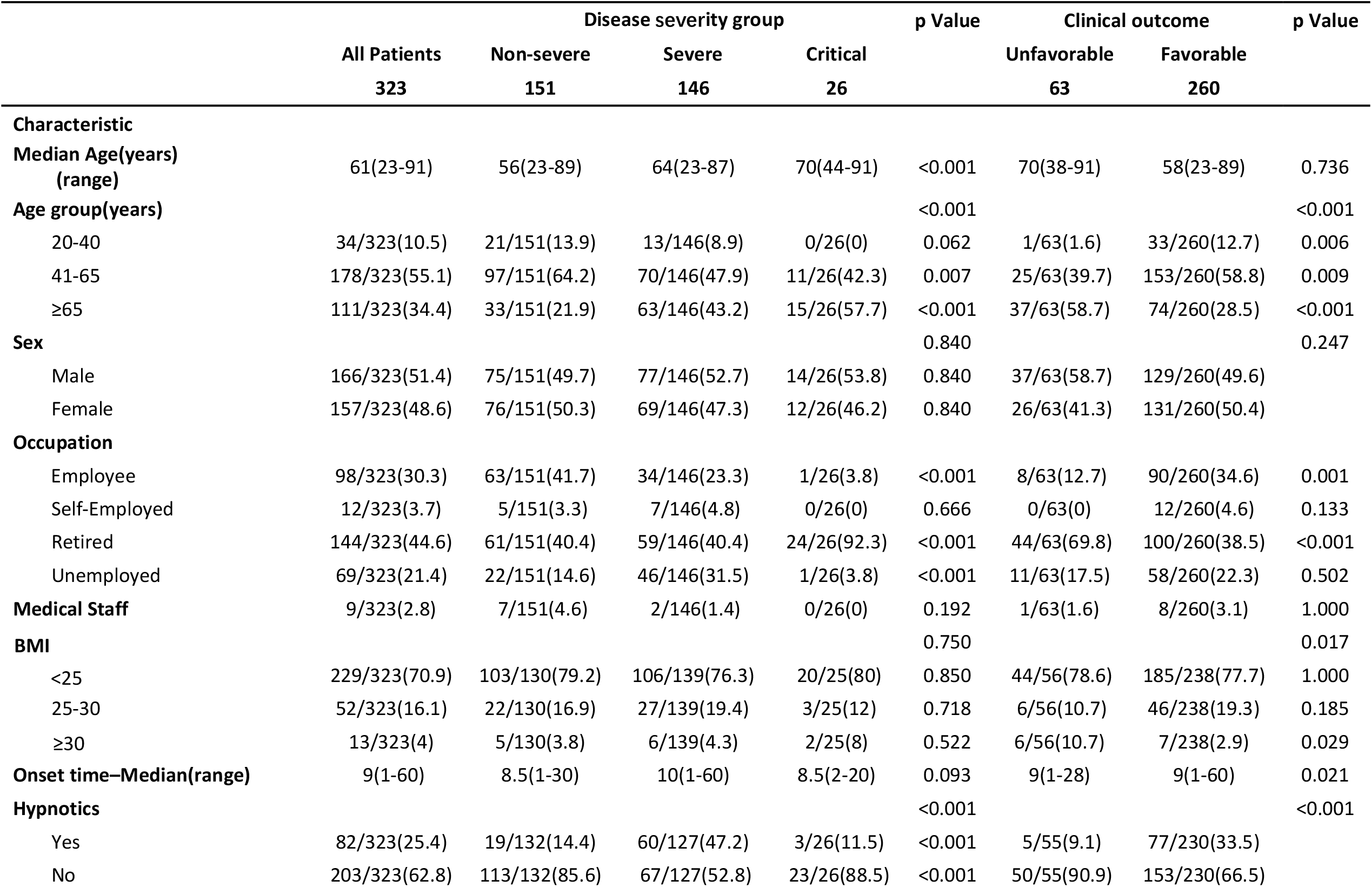
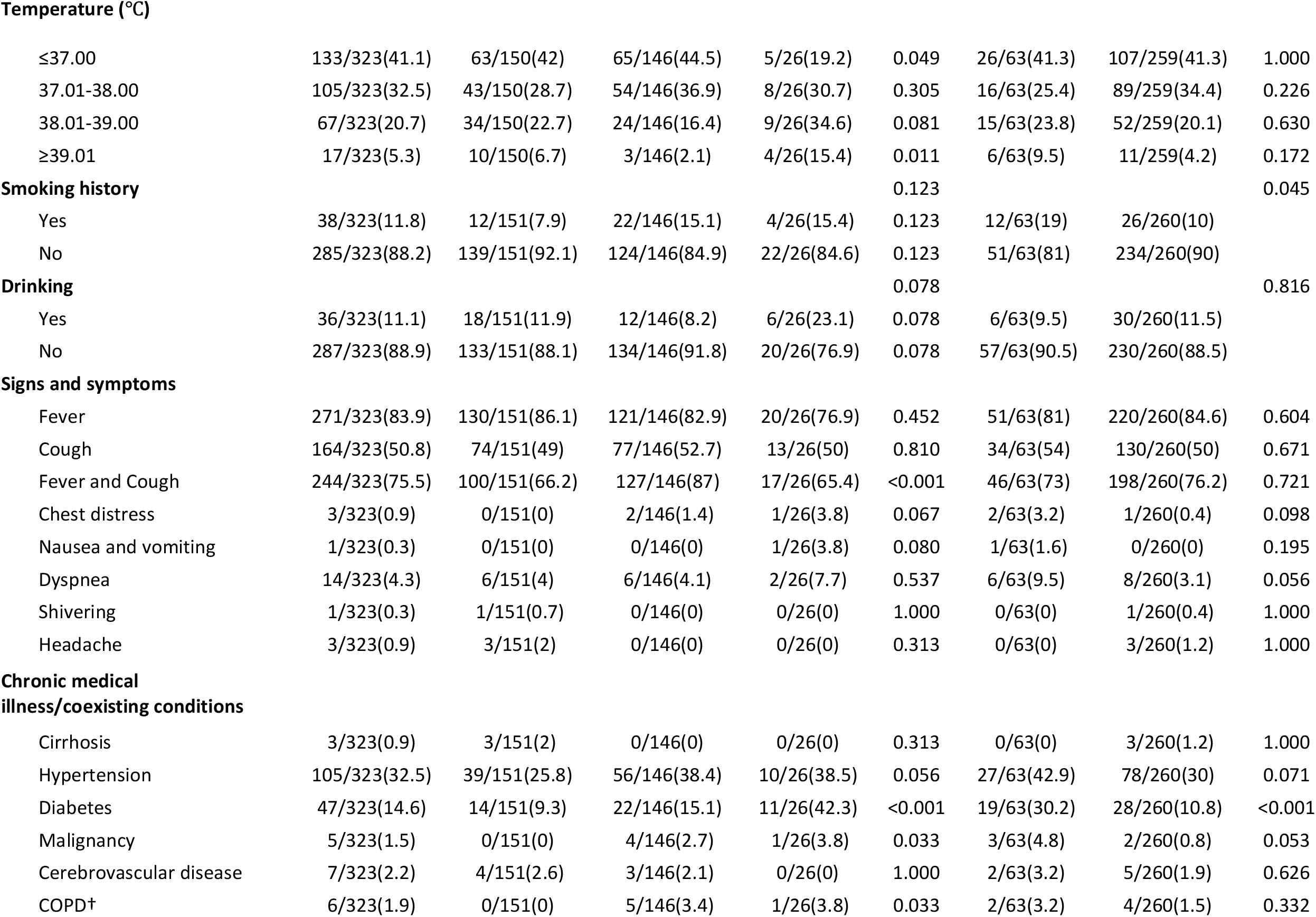
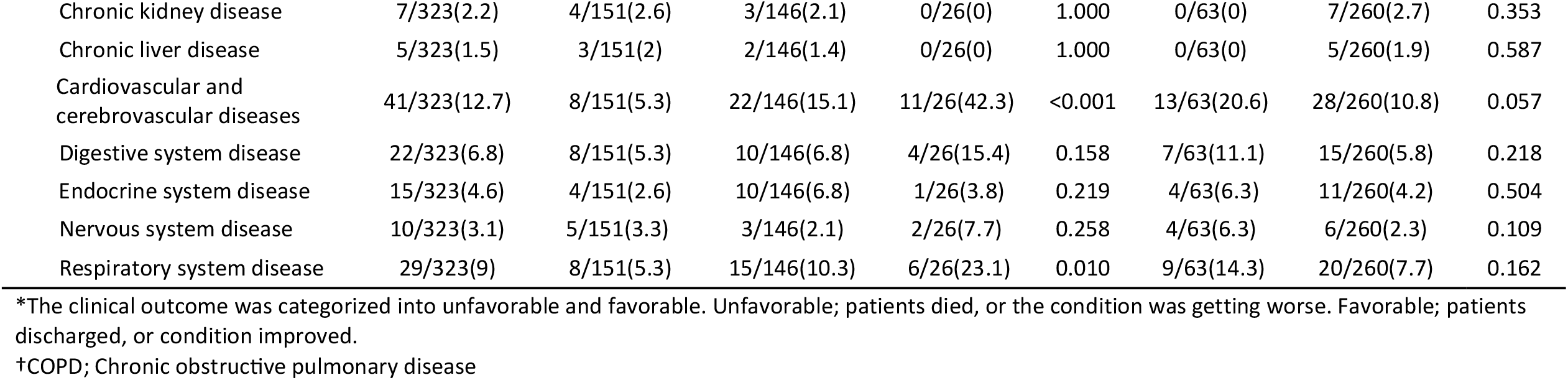
Demographics and Clinical Characteristics in disease status and clinical outcome*.

On admission, fever (83.9%, 271/323) and cough (50.8%, 164/323) were the most common symptoms, while dyspnea (4.3%, 14/323), chest distress (0.9%, 3/323) and headache (0.9%, 3/323) were uncommon.

### Clinical outcomes

The average observation period for the 323 patients was 28 days (range, 20–47 days). Favorable outcomes were recorded in 260 patients and unfavorable outcomes in 63 patients. Among the three disease severity groups, 86.8% (131/151) and 84.9% (124/146) of patients in the non-severe and severe groups, respectively, had favorable outcomes. By contrast, 80.8% (21/26) of patients in the critical group had unfavorable outcomes (Figure 1A). Patients older than 65 years showed more unfavorable than favorable outcomes. Patients with diabetes and body mass index (BMI) of ≥30 were more likely to have unfavorable outcomes (Table 1).

**Figure 1.**
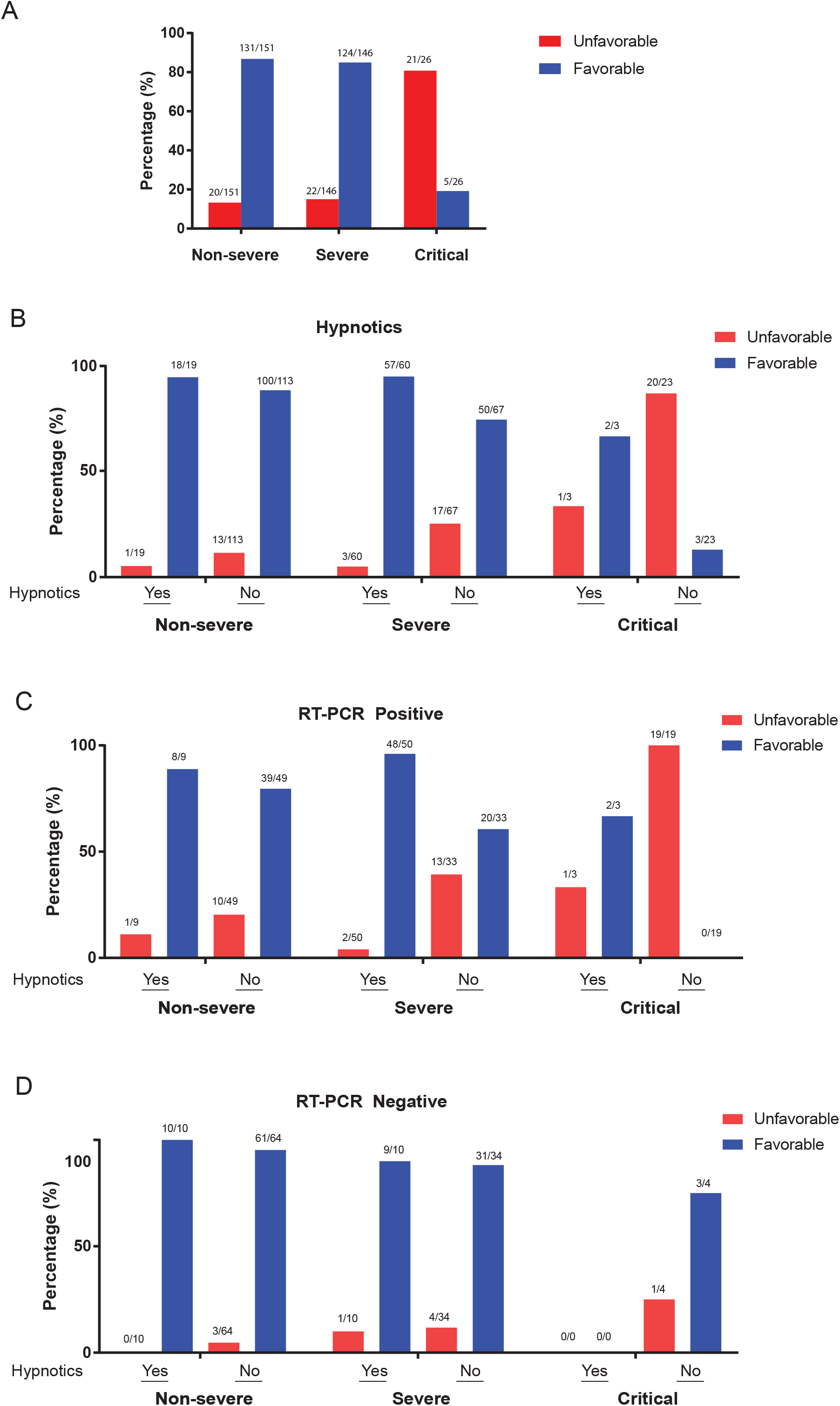
Clinical outcomes for the three disease severity groups. (A) Overall clinical outcomes of patients in the non-severe, severe, and critical disease groups. The percentages are calculated by the number of each outcome group (Unfavorable or Favorable) divided by the total number of patients in each group. (B) Clinical outcomes of patients in the non-severe, severe, and critical disease groups who were either administered hypnotics or not. (C) Clinical outcomes of RT-PCR-positive patients in the non-severe, severe, and critical disease groups who were either administered hypnotics or not. D) Clinical outcomes of RT-PCR-negative patients in the non-severe, severe, and critical disease groups who were either administered hypnotics or not. The percentages are calculated by the number of each outcome group (Unfavorable or Favorable) divided by the total number at each diagnosis status with either using (Yes) or not using (No) hypnotics.

Zopiclone, a cyclopyrrolone-class drug for insomnia, was administered at a dose of 1 mg per day to 82 patients (25.4%) for the duration of their hospitalization. Overall, favorable outcomes were recorded in 77 of these patients (Table 1). In comparing hypnotics and non-hypnotics use in patients within the three disease groups, favorable clinical outcomes were more prevalent among patients on hypnotics (94.7% vs. 88.5% for non-severe, 95% vs. 74.6% for severe, and 66.7% vs. 13.0% for critical) (*p*<0.05) (Figure 1B). And favorable clinical outcomes were associated with the administration of hypnotics among rRT-PCR-positive and rRT-PCR-negative patients in each disease severity group (Figures 1C and 1D).

### CT and laboratory abnormalities

The radiologic and laboratory test results are summarized in Table 2 (a complete version is available as Supplementary Table S1). CT abnormalities were found in 314 patients. Ground-glass opacity (GGO) findings were bilateral in 55.0% (83/151), 52.1% (76/146) and 26.9% (7/26) of patients in the non-severe, severe and critical disease groups, respectively. Multiple bilateral pulmonary consolidations and intralocular interstitial thickening were observed more frequently among patients with unfavorable outcomes (11.1%, 7/63) than those with favorable outcomes (0.8%, 2/260). Representative CT images related to clinical outcomes are shown in Figure S1.

**Table 2.**
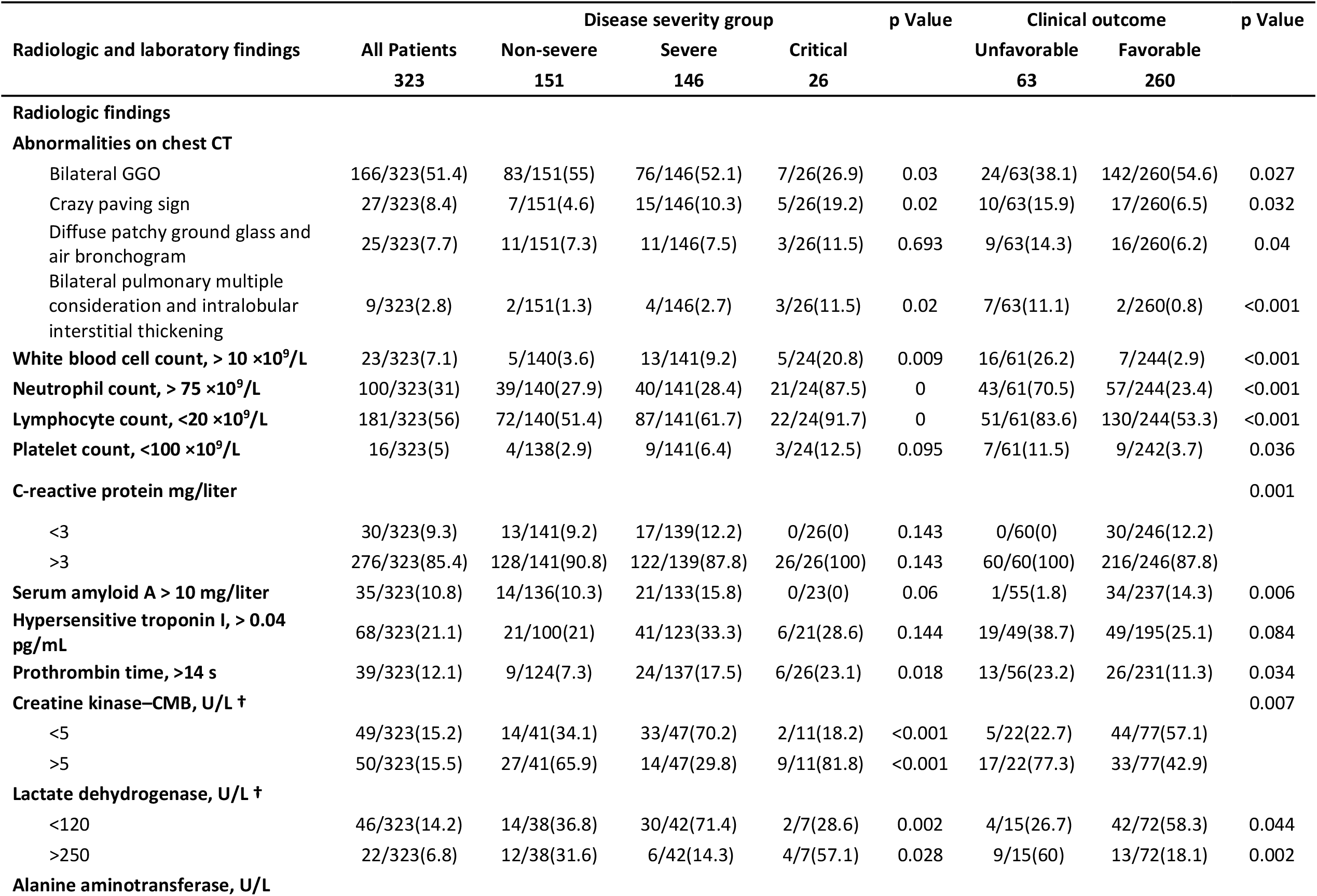
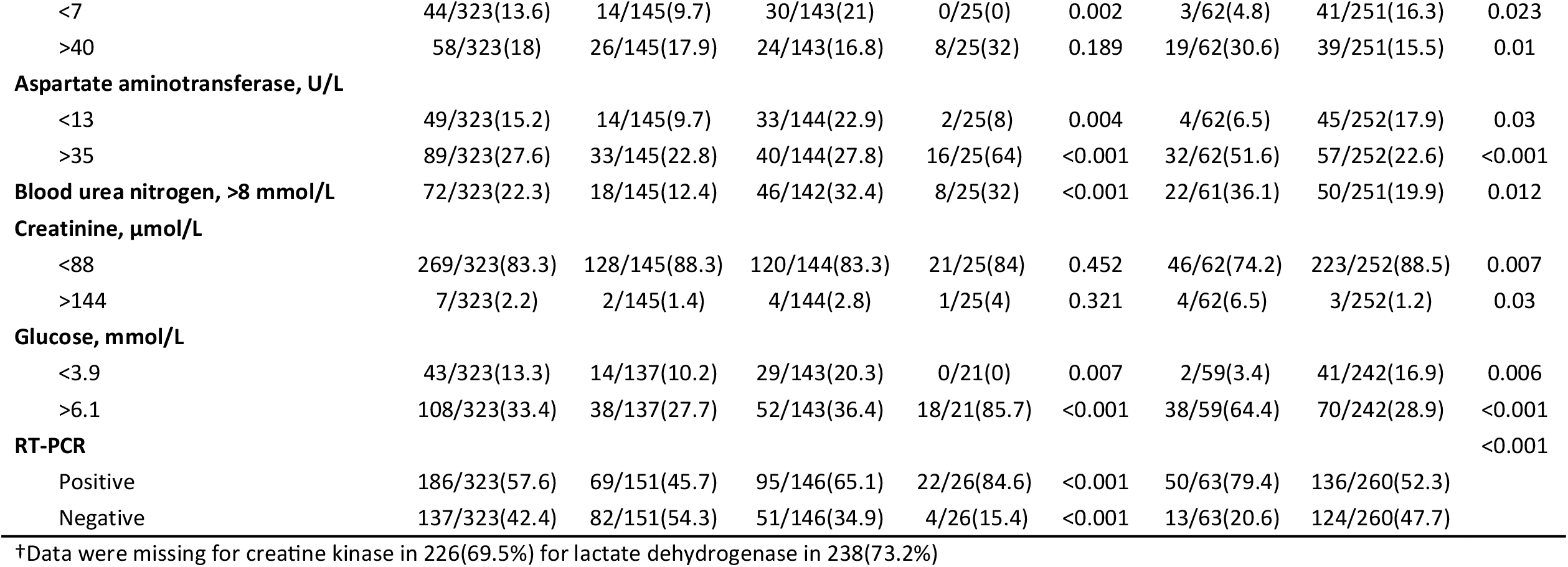
Radiographic and laboratory findings of 323 patients with COVID-19 (Part of Table S2)

Laboratory findings between patients with favorable and unfavorable outcomes showed differences in leukocyte and neutrophil counts and C-reactive protein, as well as lactate dehydrogenase, creatinine, alanine aminotransferase, aspartate aminotransferase, blood urea nitrogen, glucose, and serum amyloid A, which were all higher among patients with unfavorable outcomes. Lymphopenia also occurred among 83.6% of patients with unfavorable outcomes, while D-dimer showed no significant differences.

At the time of admission, the initial rRT-PCR was positive in 186 cases and negative in 137. rRT-PCR was positive more often among patients in the critical (84.6%) and severe (65.1%) disease groups, than in the non-severe group (45.7%). Patients whose rRT-PCR were initially negative also had better clinical outcomes, with only 9.5% (13/137) having unfavorable outcomes.

### Treatments and complications

Results related to treatment and complications are shown in Table 3. Oseltamivir (69.7%, 225/323), ganciclovir (71.2%, 230/323), and arbidol (208/323, 64.4%) were the three most frequently used antiviral medications. And one or more courses of moxifloxacin, a broad-spectrum antibiotic, was administered to 94.1% (304/323) of patients. Also, 60.7% (196/323) of patients were given corticosteroid and glucocorticoid, and 95.7% (309/323) received alternative therapy or traditional Chinese medicine. Kaletra® (lopinavir/ritonavir), an antiretroviral drug for human immunodeficiency virus infection, was more administered to patients in the critical disease group (46.2%) and in those with unfavorable than favorable outcomes (23.8% vs 5.0%). Interferon-α was also given more often to patients with unfavorable than favorable outcomes (9.5% vs. 6.2%). Other medications showed no significant differences in clinical outcomes.

**Table 3.**
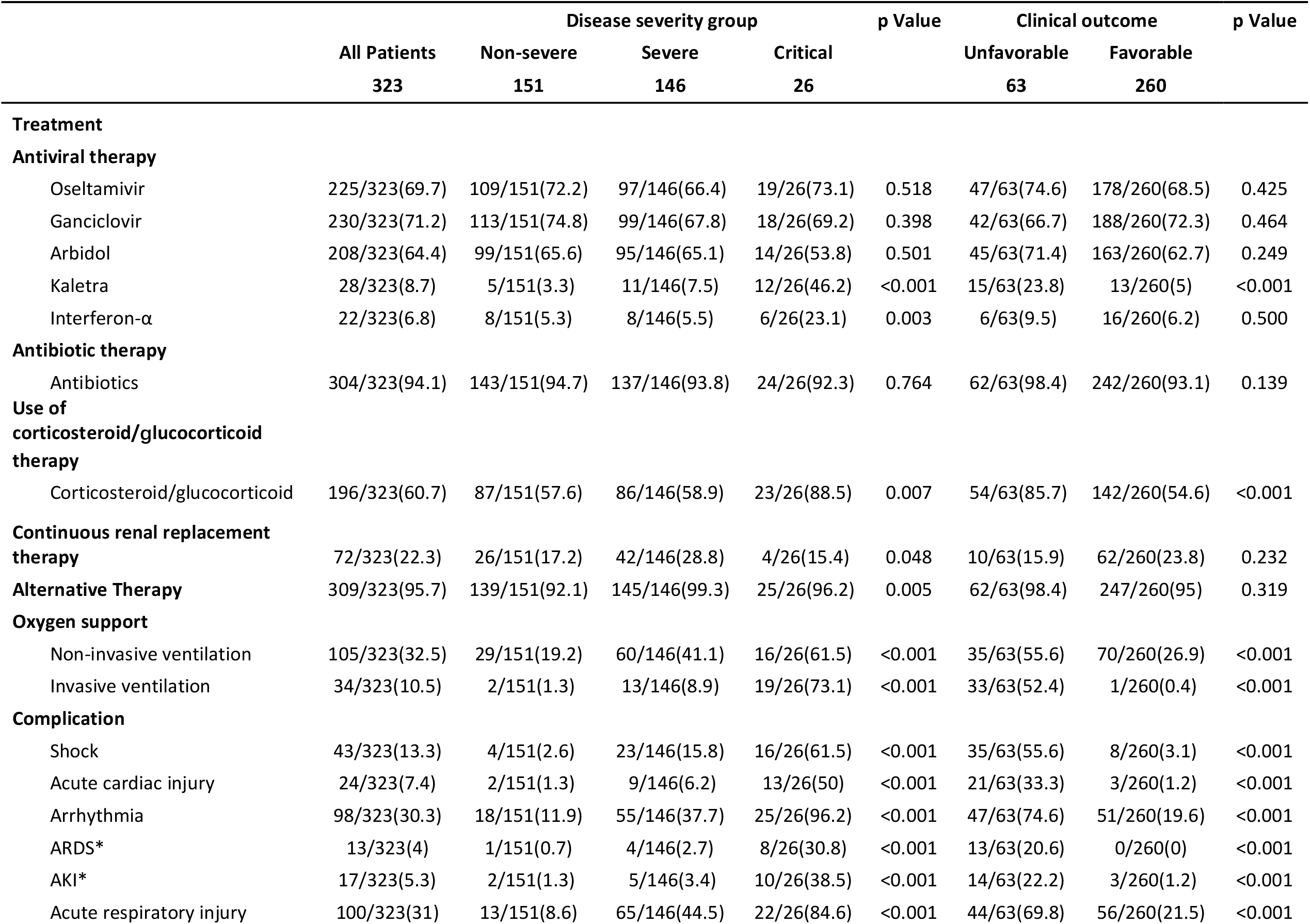
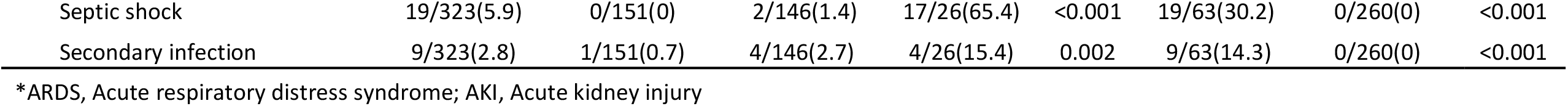
Treatments, Complications, and Clinical Outcome.

Oxygen therapy via invasive ventilation and non-invasive ventilation was also given more often to patients with unfavorable clinical outcomes. In comparing the outcome of each treatment within the non-severe or severe or critical disease groups, there was no clear improvement (Figure S2).

Of the 63 patients with unfavorable outcomes, complications, such as arrhythmia (74.6% vs. 19.6%), acute lung injury (69.8% vs. 21.5%), shock (55.6% vs. 3.1%), acute cardiac injury (33.3% vs. 1.2%), and acute respiratory distress syndrome (20.6% vs. 0%), were significantly more common in patients with favorable outcomes (Table 3).

### Risk factors associated with clinical outcomes and survival analysis

A total of 27 categorical variables were identified in univariate logistic regression analysis, namely: age, smoking, BMI, hypnotics, dyspnea, diabetes, malignancy, cardiovascular and cerebrovascular diseases, serum amyloid A, procalcitonin, hypersensitive troponin I, creatine kinase CMB, lactate dehydrogenase, alanine aminotransferase, aspartate aminotransferase, blood urea nitrogen, creatinine, glucose, leukocyte count, neutrophil count, platelet count, rRT-PCR at diagnosis, clinical status at admission, bilateral GGO, crazy paving sign, diffuse patchy ground glass and air bronchogram, and multiple bilateral pulmonary consolidation and intralobular interstitial thickening (Table S2). Eight variables were demonstrated as independent risk factors based on the multivariate logistic regression model. The results indicated that age (patients over 65 years), smoking, critical disease designation, diabetes, abnormally higher hypersensitive troponin I (>0.04 pg/mL), leukocyte count (>10×10^9^/L) and neutrophil count (>75 ×10^9^/L) were significantly associated with unfavorable clinical outcome, and hypnotics showed significant beneficial effects on clinical outcomes (*p*<0.001) (Figure 2A).

**Figure 2.**
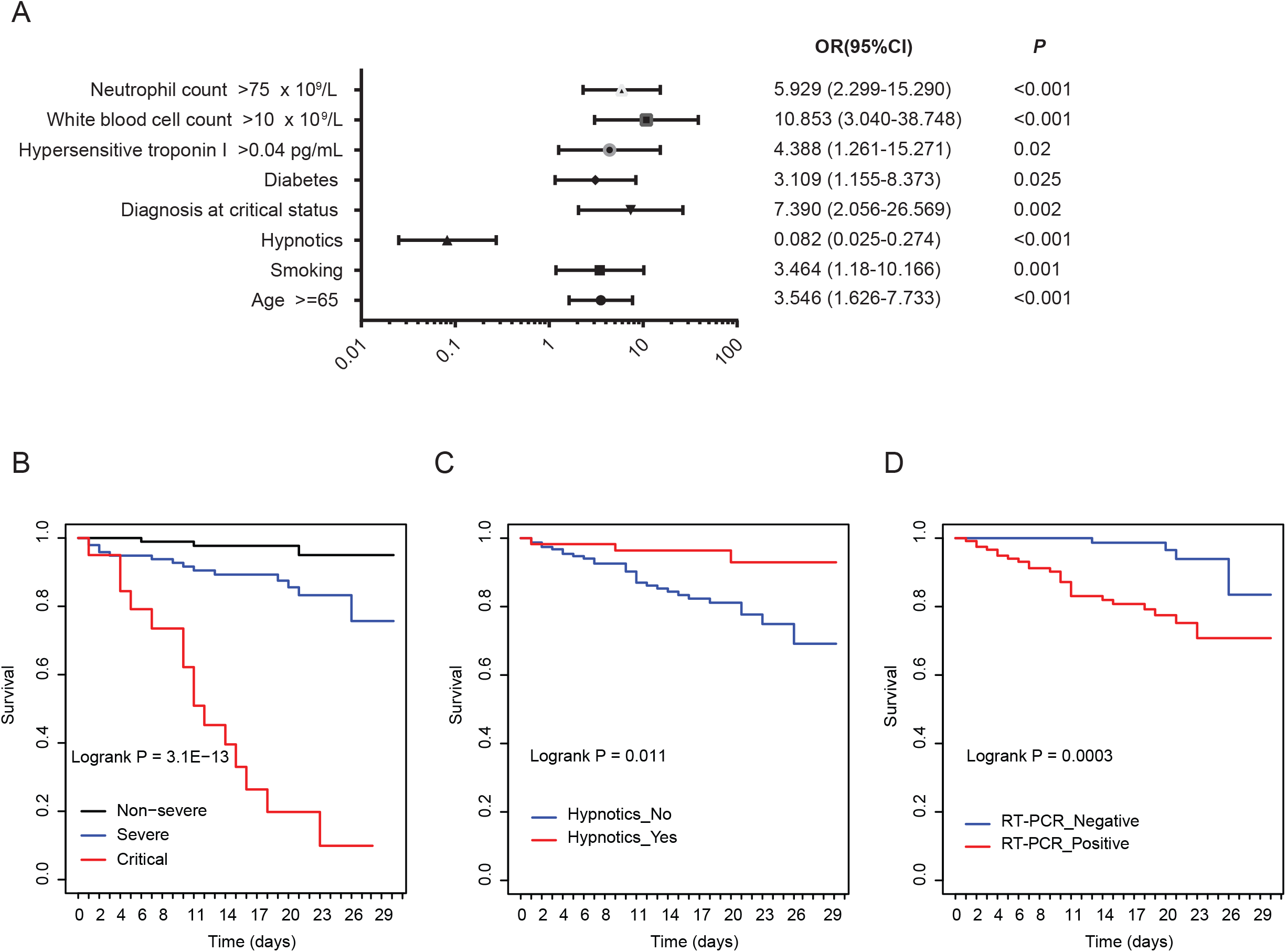
Multivariate regression and Kaplan-Meier curves of survival analysis. (A) Factors showing significantly independent association with clinical outcome. Odds ratio, 95%CI, and *P* values are derived from logistic regression modelling. (B) Kaplan-Meier curve demonstrating survival of COVID-19 patients by disease severity group: non-severe, severe, and critical. (C) Kaplan-Meier curve demonstrating survival of COVID-19 patients by the usage of hypnotics. (D) Kaplan-Meier curve demonstrating survival of COVID-19 patients by RT-PCR results. *P* values for survival analysis are derived by the log-rank test.

Patients in the non-severe group showed significantly better survival compared with those in the severe and critical groups (Figure 2B). Patients given hypnotics showed significant favorable survival compared with the non-hypnotics (Figure 2C), and patients with positive rRT-PCR results showed significantly poorer survival compared with those with negative rRT-PCR (Figure 2D).

## DISCUSSION

In contrast to a previous study,^12^ most patients with COVID-19 in the severe group of our series showed favorable clinical outcomes, at approximately the same frequency as in non-severe cases (84.9% vs 86.8%), and survival analysis demonstrated consistently higher survival rates in non-severe and severe cases than in critical cases. Although previous studies failed to show that smoking was a risk factor for COVID-19,^9,14^ multivariate analysis in our study demonstrated that smoking was an independent risk factor for unfavorable outcome. Otherwise, we confirmed findings from other studies^10,12,13^ that age over 65 years and leukocytosis with left shift were associated with poorer clinical outcome.

Zopiclone, a commonly prescribed nonbenzodiazepine soporific, was significantly associated with improved clinical outcome. Patients taking hypnotics showed better outcome at the same disease stage than those not taking hypnotics. For patients in the more severe disease groups, the improvement effect was even more pronounced. Also, in analyzing the effect of hypnotics on rRT-PCR-positive and rRT-PCR-negative patients separately, we found hypnotics had more striking effect on the former. Moreover, hypnotics were identified as an independent factor in the risk model that contributed to better clinical outcomes. Also, patients administered hypnotics had a better survival rate.

Patients usually showed strong anxiety, sleep deficiency and oxygen insufficiency during disease progression, which will lead to the metabolic dysregulation^18,19^ and immune system abnormalities^20^. Better sleep quality and stress reduction could be one partial reason for prescribing hypnotics to COVID-19 patients. Dimitrov et al indicated that sleep can exert some immune-supportive effects and potentially enhance an effective T cell response, and it has high relevant with some specific sleep disorders or impaired sleep, such as depression and chronic stress.^21^

In addition, the superior efficacy of zopiclone may be due to enhanced gamma aminobutyric acid (GABA) signaling. That is, zopiclone can interact with GABA^A^, and GABA^A^ receptor can magnify responses to GABA.^22^ GABA signaling promotes autophagy activation, which improves phagosome maturation and promotes host protection against infections.^23^ To our knowledge, the beneficial effect of hypnotics on clinical outcome has not been reported previously in the management of COVID-19 patients.

Up till now, several description studies have mentioned the non-effective function of current medication treatment no matter which stages or outcomes was^10,12,24^, and it was accord with our study analysis. By specifically comparing the standard treatment effect at the same disease status for each treatment (Figure S2), we didn’t see any standard therapy could improve clinical outcome in the study. However, since only about 25% of patients in our study took hypnotics, we assume that self-healing could be the major reason for the high recovery rate of patients in the non-severe and severe disease groups. That is, COVID-19 is most likely a self-limited disease in the majority of patients.

Both rRT-PCR-confirmed COVID-19 patients and clinically diagnosed patients who were rRT-PCR negative were included in this study. Due to the burgeoning epidemic and high exposure situation in Wuhan, the guidelines for COVID-19 diagnosis and treatment indicated that residents of Wuhan with clinical presentations suggestive of COVID-19 (including respiratory symptoms, CT scan results and laboratory tests excluding other infectious causes of pneumonia) could be admitted to hospital irrespective of the rRT-PCR result. Actually, all of the rRT-PCR-negative patients had CT image features compatible with COVID-19. Moreover, the high false-negativity of rRT-PCR (about 20-40%)^25^ presents a significant burden on health care providers to use their clinical judgment. And chest CT has a higher sensitivity for diagnosis of COVID-19 than rRT-PCR.^25^

Several underlying reasons including uneven quality from different detection kits, improper collection of throat swab specimens, and low concentration of virus in samples^26^ can lead to the possible results deviation. Therefore, including the rRT-PCR-negative patients was an important measure to control and prevent the spread of COVID-19 in Wuhan. We found that patients in the severe or critical disease groups were more likely to be rRT-PCR positive. Survival analysis also corroborated that rRT-PCR-positive patients showed poorer clinical outcomes.

Since all patients in our study were tested by the same experienced team using the same rRT-PCR protocol, we believe the major reason for the negative rRT-PCR results in our patient series was due to the low concentration of virus in the throat, which may indicate that patients with negative rRT-PCR test might be less likely to infect other people. We found that only a small portion of patients (less than 10%) with negative rRT-PCR had unfavorable outcomes. In performing follow-up rRT-PCR on patients in the severe disease group with abnormal CT, we found 23 cases whose first test was negative and later tests were positive. We believe the inclusion of rRT-PCR-negative patients with clinically compatible presentation of COVID-19 will help guide clinicians in the care of such patients.

Higher BMI (≥30), hyperglycemia and diabetes, and cardiovascular disease were distinct risk factors for unfavorable clinical outcomes. Angiotensin-converting enzyme-2 (ACE2), which serves as a cell-entry receptor for SARS CoV-2,^27^ plays a protective role for both diabetes and cardiovascular diseases.^28,29^ Kuba and colleagues demonstrated that SARS CoV downregulates ACE2 protein,^30^ which could explain why COVID-19 patients with diabetes and cardiovascular disease have worse clinical outcomes.

We found that abnormally high hypersensitive troponin I was an independent predictor for poor clinical outcome. Increased troponin can enhance coagulation activation.^31^ In patients with COVID-19, immune damage to the hematopoietic system, ischemic hypoxia-reperfusion injury, and drugs can cause coagulation disorders.^8,31-34^ We speculate that increased troponin will induce dysfunction of coagulation and thrombus formation with possible pulmonary embolism, which would further aggravate the patient’s condition.

There were some limitations in our study. Incomplete laboratory test results in some patient records may have caused deviations in statistical analysis. Except for hypnotics, we found that all treatments were ineffective and many treatments showed unwanted side effects, including liver injury.^35-37^ Although we did not conduct separate analysis for rRT-PCR-positive and rRT-PCR-negative patients, our multivariate analysis identified eight independent risk factors, which were independent of the rRT-PCR result.

Although the vast majority of patients recovered, approximately 20% of our hospitalized patient cohort had unfavorable clinical outcomes. To what extent chronic respiratory insufficiency or other organ system sequelae occur in COVID-19 patients will require careful and prolonged follow-up studies. So far, it seems there is no effective standard treatment. However, we have found that using hypnotics could significantly improve clinical outcome of COVID-19. We also found that some novel risk factors that could predict patient outcome, which can help in early decision making for improving treatment outcomes of COVID-19 patients.

## Data Availability

Data sharing not applicable

## ACKNOWLEDGEMENTS

We thank all of the staff members of the Tianyou Hospital, affiliated to Wuhan University of Science and Technology (Li Ming Liu, Jing Zhang, Qiong Luo, Guilian Shang, Ting Li, Qiushi Zhang, Shuan Liu) for their efforts in collecting the information used in this study.

---

**Figure S1.**
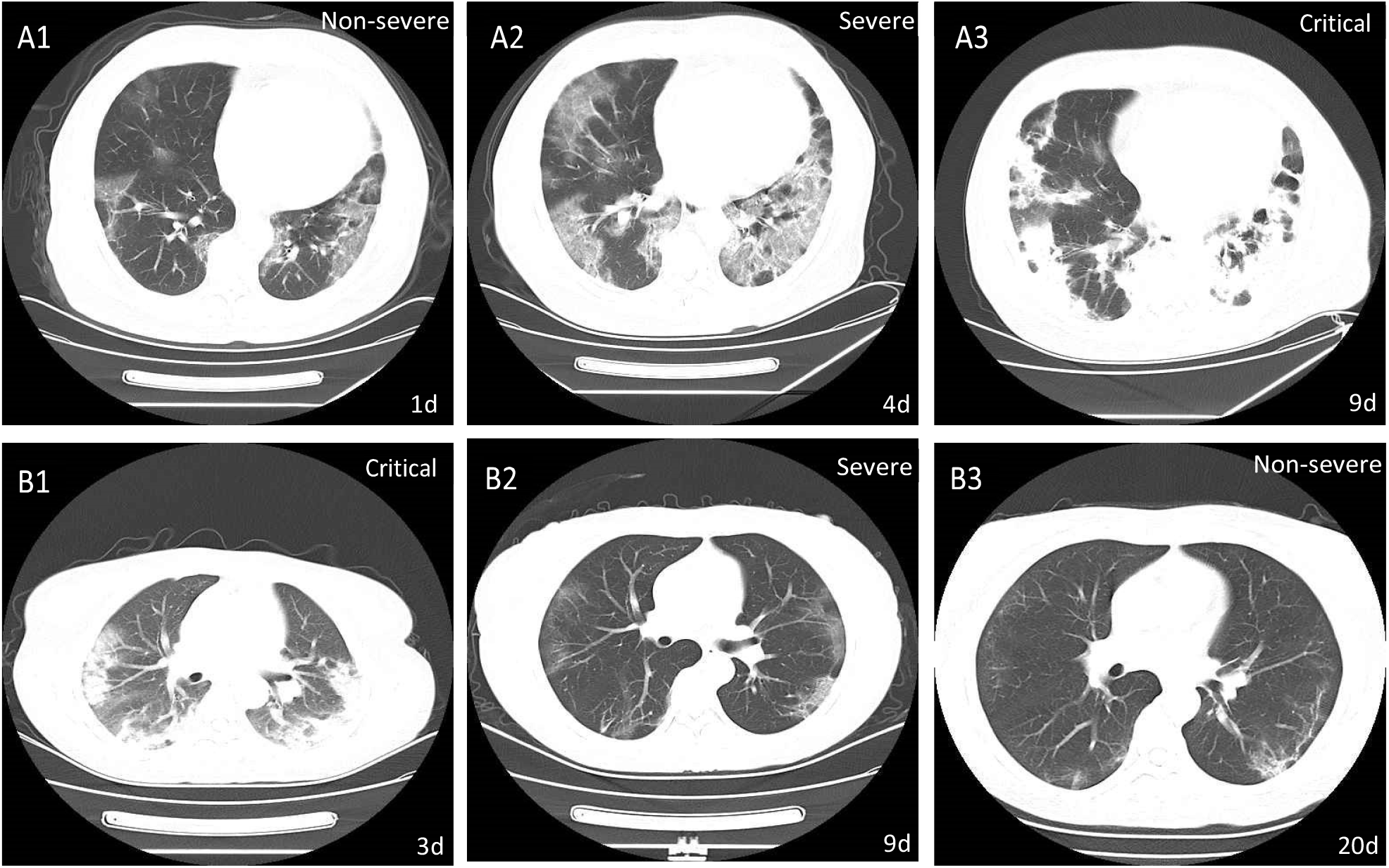
Representative dynamic changes in chest computer tomography (CT) scans from two patients with COVID-19. A1–A3, CT images of a 70-year-old male patient, who progressed from the non-severe to critical disease group, and died. A1 (day 1), A2 (day 4), A3 (day 9): below the pleura are scattered shadows of frosted glass, large sheet diffuse paving stones in both lungs, and extensive consolidation of both lungs with thickening of interlobular stroma repetitively. B1–B3, CT images of a 44-year-old female patient, who progressed from the critical to non-severe disease group. B1 (day 3) exhibiting extensive consolidation of both lungs; B2 (day 9) exhibiting frosted hyaline change and paving stone signs. B3 (day 20) showing significant resolution of ground-glass opacities and subpleural cord changes.

**Figure S2:**
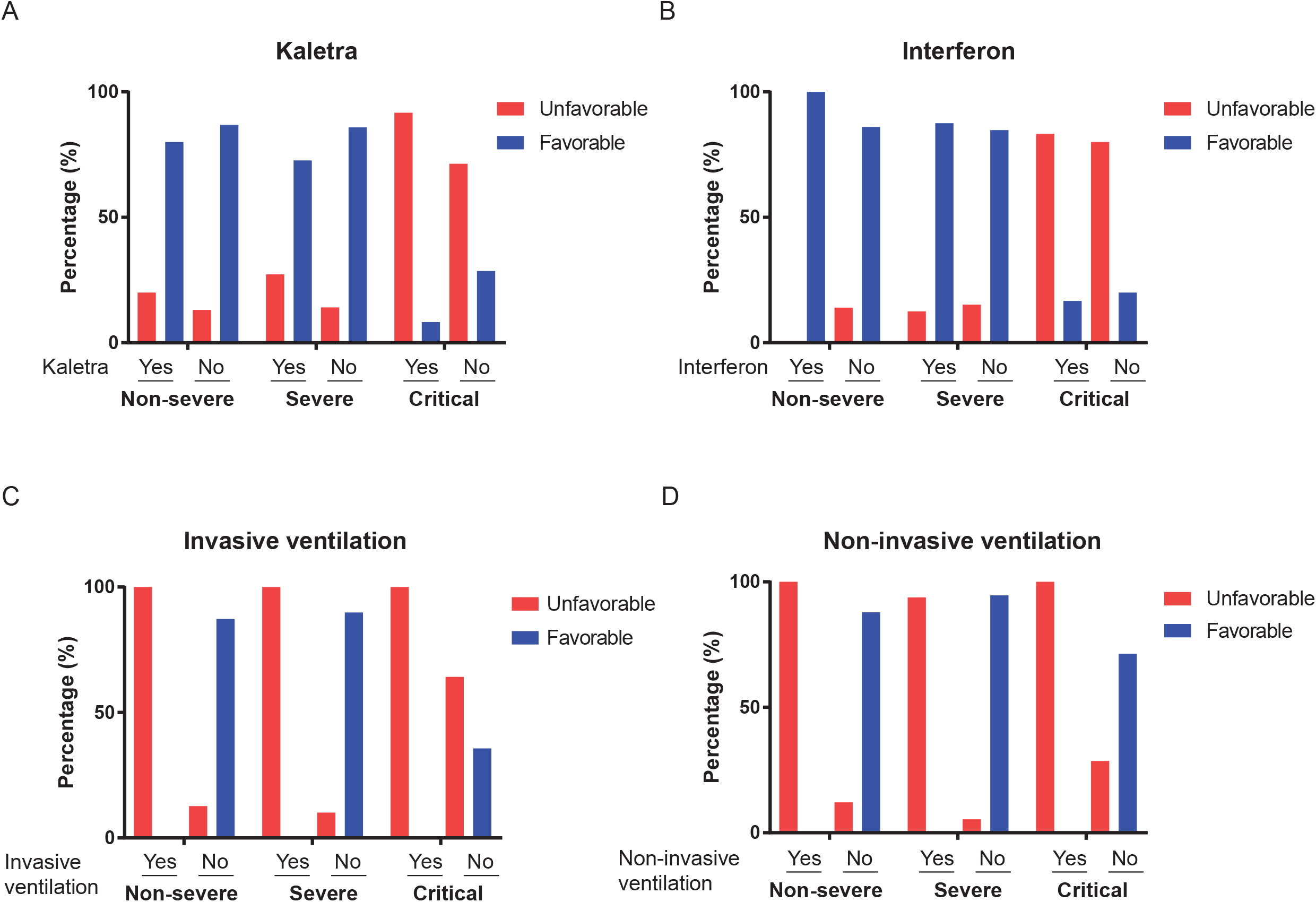
The association of different treatments with clinical outcome in three diagnostic status. The association of clinical outcome with three diagnosis status: Non-severe, Severe, and Critical under the treatment A) Kaletra, B) Interferon, C) Invasive ventilation, D) Non-invasive ventilation. The percentages are calculated by the number of each outcome group (Unfavorable or Favorable) divided by the total number at each diagnosis status with (Yes) using or not (No) using a treatment.

**Supplementary Table S1.** Radiographic and laboratory findings of 323 patients with COVID-19.

| Radiologic and laboratory findings | Disease severity group |  |  |  | p Value | Clinical outcome |  | p Value |
| --- | --- | --- | --- | --- | --- | --- | --- | --- |
|  | All Patients | Non-severe | Severe | Critical |  | Unfavorable | Favorable |  |
|  | 323 | 151 | 146 | 26 |  | 63 | 260 |  |
| Radiologic findings |  |  |  |  |  |  |  |  |
| Abnormalities on chest CT |  |  |  |  |  |  |  |  |
| No GGO | 9/323(2.8) | 4/151(2.6) | 4/146(2.7) | 1/26(3.8) | 0.768 | 0/63(0) | 9/260(3.5) | 0.214 |
| Local GGO | 17/323(5.3) | 11/151(7.3) | 6/146(4.1) | 0/26(0) | 0.317 | 1/63(1.6) | 16/260(6.2) | 0.211 |
| Bilateral GGO | 166/323(51.4) | 83/151(55) | 76/146(52.1) | 7/26(26.9) | 0.030 | 24/63(38.1) | 142/260(54.6) | 0.027 |
| Combination of patchy ground glass opacity and pulmonary consolidation | 61/323(18.9) | 28/151(18.5) | 29/146(19.9) | 4/26(15.4) | 0.930 | 8/63(12.7) | 53/260(20.4) | 0.223 |
| Crazy paving sign | 27/323(8.4) | 7/151(4.6) | 15/146(10.3) | 5/26(19.2) | 0.020 | 10/63(15.9) | 17/260(6.5) | 0.032 |
| Diffuse patchy ground glass and air bronchogram | 25/323(7.7) | 11/151(7.3) | 11/146(7.5) | 3/26(11.5) | 0.693 | 9/63(14.3) | 16/260(6.2) | 0.040 |
| Bilateral pulmonary multiple consideration and intralobular interstitial thickening | 9/323(2.8) | 2/151(1.3) | 4/146(2.7) | 3/26(11.5) | 0.020 | 7/63(11.1) | 2/260(0.8) | <0.001 |
| Laboratory findings |  |  |  |  |  |  |  |  |
| PH* |  |  |  |  | 0.036 |  |  | 0.478 |
| <7.35 | 1/323(0.3) | 0/17(0) | 0/33(0) | 1/11(9.1) | 0.180 | 1/19(5.3) | 0/42(0) | 0.311 |
| 7.35-7.45 | 20/323(6.2) | 9/17(52.9) | 7/33(21.2) | 4/11(36.4) | 0.071 | 6/19(31.6) | 14/42(33.3) | 1.000 |
| >7.45 | 40/323(12.4) | 8/17(47.1) | 26/33(78.8) | 6/11(54.5) | 0.062 | 12/19(63.2) | 28/42(66.7) | 1.000 |
| **Lactate, mmol/L \*** |  |  |  |  | 0.290 |  |  | 0.533 |
| <0.5 | 1/323(0.3) | 0/15(0) | 1/33(3) | 0/11(0) | 1.000 | 0/18(0) | 1/41(2.4) | 1.000 |
| 0.5-1.6 | 44/323(13.6) | 13/15(86.7) | 25/33(75.8) | 6/11(54.5) | 0.176 | 12/18(66.7) | 32/41(78) | 0.549 |
| >1.6 | 14/323(4.3) | 2/15(13.3) | 7/33(21.2) | 5/11(45.5) | 0.169 | 6/18(33.3) | 8/41(19.5) | 0.414 |
| **PaO2, mmHg \*** |  |  |  |  | 0.943 |  |  | 0.593 |
| <80 | 36/323(11.1) | 11/17(64.7) | 19/33(57.6) | 6/11(54.5) | 0.882 | 11/19(57.9) | 25/42(59.5) | 1.000 |
| 80-100 | 12/323(3.7) | 3/17(17.6) | 6/33(18.2) | 3/11(27.3) | 0.825 | 5/19(26.3) | 7/42(16.7) | 0.489 |
| >100 | 13/323(4) | 3/17(17.6) | 8/33(24.2) | 2/11(18.2) | 0.915 | 3/19(15.8) | 10/42(23.8) | 0.737 |
| **PaCO2, mmHg \*** |  |  |  |  | 0.006 |  |  | 1.000 |
| <35 | 36/323(11.1) | 10/17(58.8) | 24/33(72.7) | 2/11(18.2) | 0.006 | 11/19(57.9) | 25/42(59.5) | 1.000 |
| 35-45 | 25/323(7.7) | 7/17(41.2) | 9/33(27.3) | 9/11(81.8) | 0.006 | 8/19(42.1) | 17/42(40.5) | 1.000 |
| **PaO2/Fio2\*** |  |  |  |  | 0.301 |  |  | 0.422 |
| <400 | 48/323(14.9) | 11/16(68.8) | 28/32(87.5) | 9/11(81.8) | 0.292 | 16/18(88.9) | 32/41(78) | 0.476 |
| 400-500 | 6/323(1.9) | 2/16(12.5) | 2/32(6.2) | 2/11(18.2) | 0.522 | 2/18(11.1) | 4/41(9.8) | 1.000 |
| >500 | 5/323(1.5) | 3/16(18.8) | 2/32(6.2) | 0/11(0) | 0.295 | 0/18(0) | 5/41(12.2) | 0.310 |
| **White blood cell count, ×10<sup>9</sup>/L** |  |  |  |  | 0.041 |  |  | <0.001 |
| <4 | 90/323(27.9) | 46/140(32.9) | 39/141(27.7) | 5/24(20.8) | 0.444 | 14/61(23) | 76/244(31.1) | 0.272 |
| 4-10 | 192/323(59.4) | 89/140(63.6) | 89/141(63.1) | 14/24(58.3) | 0.885 | 31/61(50.8) | 161/244(66) | 0.041 |
| >10 | 23/323(7.1) | 5/140(3.6) | 13/141(9.2) | 5/24(20.8) | 0.009 | 16/61(26.2) | 7/244(2.9) | <0.001 |
| **Neutrophil count, ×10<sup>9</sup>/L** |  |  |  |  | 0.000 |  |  | <0.001 |
| <40 | 70/323(21.7) | 28/140(20) | 42/141(29.8) | 0/24(0) | 0.001 | 8/61(13.1) | 62/244(25.4) | 0.061 |
| 40-75 | 135/323(41.8) | 73/140(52.1) | 59/141(41.8) | 3/24(12.5) | 0.001 | 10/61(16.4) | 125/244(51.2) | <0.001 |
| >75 | 100/323(31) | 39/140(27.9) | 40/141(28.4) | 21/24(87.5) | <0.001 | 43/61(70.5) | 57/244(23.4) | <0.001 |
| **Lymphocyte count, ×10<sup>9</sup>/L** |  |  |  |  | 0.002 |  |  | <0.001 |
| <20 | 181/323(56) | 72/140(51.4) | 87/141(61.7) | 22/24(91.7) | 0.000 | 51/61(83.6) | 130/244(53.3) | <0.001 |
| 20-50 | 120/323(37.2) | 66/140(47.1) | 52/141(36.9) | 2/24(8.3) | 0.001 | 10/61(16.4) | 110/244(45.1) | <0.001 |
| >50 | 4/323(1.2) | 2/140(1.4) | 2/141(1.4) | 0/24(0) | 1.000 | 0/61(0) | 4/244(1.6) | 0.587 |
| **Monocyte count, ×10<sup>9</sup>/L** |  |  |  |  | 0.305 |  |  | 0.127 |
| <3 | 103/323(31.9) | 44/138(31.9) | 54/141(38.3) | 5/24(20.8) | 0.195 | 27/61(44.3) | 76/242(31.4) | 0.081 |
| 3-10 | 159/323(49.2) | 72/138(52.2) | 70/141(49.6) | 17/24(70.8) | 0.157 | 29/61(47.5) | 130/242(53.7) | 0.471 |
| >10 | 41/323(12.7) | 22/138(15.9) | 17/141(12.1) | 2/24(8.3) | 0.576 | 5/61(8.2) | 36/242(14.9) | 0.212 |
| **Platelet count, ×10<sup>9</sup>/L** |  |  |  |  | 0.154 |  |  | 0.014 |
| <100 | 16/323(5) | 4/138(2.9) | 9/141(6.4) | 3/24(12.5) | 0.095 | 7/61(11.5) | 9/242(3.7) | 0.036 |
| 100-300 | 232/323(71.8) | 112/138(81.2) | 102/141(72.3) | 18/24(75) | 0.217 | 48/61(78.7) | 184/242(76) | 0.788 |
| >300 | 55/323(17) | 22/138(15.9) | 30/141(21.3) | 3/24(12.5) | 0.433 | 6/61(9.8) | 49/242(20.2) | 0.089 |
| **C-reactive protein level mg/liter** |  |  |  |  | 0.143 |  |  | 0.001 |
| <3 | 30/323(9.3) | 13/141(9.2) | 17/139(12.2) | 0/26(0) | 0.143 | 0/60(0) | 30/246(12.2) |  |
| >3 | 276/323(85.4) | 128/141(90.8) | 122/139(87.8) | 26/26(100) | 0.143 | 60/60(100) | 216/246(87.8) |  |
| **SAA mg/liter** |  |  |  |  | 0.061 |  |  | 0.006 |
| <10 | 35/323(10.8) | 14/136(10.3) | 21/133(15.8) | 0/23(0) | 0.060 | 1/55(1.8) | 34/237(14.3) |  |
| >10 | 256/323(79.3) | 121/136(89) | 112/133(84.2) | 23/23(100) | 0.071 | 54/55(98.2) | 202/237(85.2) |  |
| **Prothrombin time, s** |  |  |  |  | 0.713 |  |  | 0.044 |
| <9 | 18/323(5.6) | 6/124(4.8) | 10/137(7.3) | 2/26(7.7) | 0.663 | 1/56(1.8) | 17/231(7.4) | 0.214 |
| >14 | 39/323(12.1) | 9/124(7.3) | 24/137(17.5) | 6/26(23.1) | 0.018 | 13/56(23.2) | 26/231(11.3) | 0.034 |
| **Activated partial thromboplastin time, s** | 30.7(0-2406) | 33(0.014-57) | 34.3(0-59.3) | 26.9(1-2406) | 0.001 | 34.2(1.193-57) | 30.4(0-2406) | 0.098 |
| **D-dimer mg/liter** |  |  |  |  | 0.002 |  |  | 0.137 |
| <0.5 | 160/323(49.5) | 83/120(69.2) | 68/138(49.3) | 9/21(42.9) | 0.002 | 24/52(46.2) | 136/228(59.6) |  |
| >0.5 | 119/323(36.8) | 37/120(30.8) | 70/138(50.7) | 12/21(57.1) | 0.003 | 27/52(51.9) | 92/228(40.4) |  |
| **Hypersensitive troponin I, pg/mL** |  |  |  |  | 0.124 |  |  | 0.084 |
| <0.04 | 176/323(54.5) | 79/100(79) | 82/123(66.7) | 15/21(71.4) | 0.086 | 30/49(61.2) | 146/195(74.9) |  |
| >0.04 | 68/323(21.1) | 21/100(21) | 41/123(33.3) | 6/21(28.6) | 0.144 | 19/49(38.7) | 49/195(25.1) |  |
| **Creatine kinase–CMB, U/L †** |  |  |  |  | <0.001 |  |  | 0.007 |
| <5 | 49/323(15.2) | 14/41(34.1) | 33/47(70.2) | 2/11(18.2) | <0.001 | 5/22(22.7) | 44/77(57.1) |  |
| >5 | 50/323(15.5) | 27/41(65.9) | 14/47(29.8) | 9/11(81.8) | <0.001 | 17/22(77.3) | 33/77(42.9) |  |
| **Lactate dehydrogenase, U/L †** |  |  |  |  | 0.007 |  |  | 0.004 |
| <120 | 46/323(14.2) | 14/38(36.8) | 30/42(71.4) | 2/7(28.6) | 0.002 | 4/15(26.7) | 42/72(58.3) | 0.044 |
| 120 - 250 | 19/323(5.9) | 12/38(31.6) | 6/42(14.3) | 1/7(14.3) | 0.159 | 2/15(13.3) | 17/72(23.6) | 0.506 |
| >250 | 22/323(6.8) | 12/38(31.6) | 6/42(14.3) | 4/7(57.1) | 0.028 | 9/15(60) | 13/72(18.1) | 0.002 |
| **Alanine aminotransferase, U/L** |  |  |  |  | 0.005 |  |  | 0.004 |
| <7 | 44/323(13.6) | 14/145(9.7) | 30/143(21) | 0/25(0) | 0.002 | 3/62(4.8) | 41/251(16.3) | 0.023 |
| 7-40 | 211/323(65.3) | 105/145(72.4) | 89/143(62.2) | 17/25(68) | 0.183 | 40/62(64.5) | 171/251(68.1) | 0.695 |
| >40 | 58/323(18) | 26/145(17.9) | 24/143(16.8) | 8/25(32) | 0.189 | 19/62(30.6) | 39/251(15.5) | 0.010 |
| **Aspartate aminotransferase, U/L** |  |  |  |  | <0.001 |  |  | <0.001 |
| <13 | 49/323(15.2) | 14/145(9.7) | 33/144(22.9) | 2/25(8) | 0.004 | 4/62(6.5) | 45/252(17.9) | 0.030 |
| 13-35 | 176/323(54.5) | 98/145(67.6) | 71/144(49.3) | 7/25(28) | <0.001 | 26/62(41.9) | 150/252(59.5) | 0.018 |
| >35 | 89/323(27.6) | 33/145(22.8) | 40/144(27.8) | 16/25(64) | <0.001 | 32/62(51.6) | 57/252(22.6) | <0.001 |
| **Total bilirubin, mmol/L** |  |  |  |  | 0.076 |  |  | 0.152 |
| <5 | 21/323(6.5) | 10/145(6.9) | 11/144(7.6) | 0/25(0) | 0.511 | 1/62(1.6) | 20/252(7.9) | 0.090 |
| 5-22 | 248/323(76.8) | 121/145(83.4) | 109/144(75.7) | 18/25(72) | 0.181 | 50/62(80.6) | 198/252(78.6) | 0.853 |
| >22 | 45/323(13.9) | 14/145(9.7) | 24/144(16.7) | 7/25(28) | 0.030 | 11/62(17.7) | 34/252(13.5) | 0.514 |
| **Blood urea nitrogen, mmol/L** |  |  |  |  | 0.001 |  |  | 0.018 |
| <3 | 51/323(15.8) | 26/145(17.9) | 21/142(14.8) | 4/25(16) | 0.776 | 6/61(9.8) | 45/251(17.9) | 0.180 |
| 3-8 | 189/323(58.5) | 101/145(69.7) | 75/142(52.8) | 13/25(52) | 0.009 | 33/61(54.1) | 156/251(62.2) | 0.313 |
| >8 | 72/323(22.3) | 18/145(12.4) | 46/142(32.4) | 8/25(32) | <0.001 | 22/61(36.1) | 50/251(19.9) | 0.012 |
| **Creatinine, µmol/L** |  |  |  |  | 0.587 |  |  | 0.007 |
| <88 | 269/323(83.3) | 128/145(88.3) | 120/144(83.3) | 21/25(84) | 0.452 | 46/62(74.2) | 223/252(88.5) | 0.007 |
| 88-144 | 38/323(11.8) | 15/145(10.3) | 20/144(13.9) | 3/25(12) | 0.623 | 12/62(19.4) | 26/252(10.3) | 0.082 |
| >144 | 7/323(2.2) | 2/145(1.4) | 4/144(2.8) | 1/25(4) | 0.321 | 4/62(6.5) | 3/252(1.2) | 0.030 |
| **Procalcitonin, ng/mL** |  |  |  |  | 0.018 |  |  | 0.624 |
| <0.1 | 198/323(61.3) | 98/117(83.8) | 88/123(71.5) | 12/20(60) | 0.018 | 37/51(72.5) | 161/209(77) |  |
| >0.1 | 62/323(19.2) | 19/117(16.2) | 35/123(28.5) | 8/20(40) | 0.018 | 14/51(27.5) | 48/209(23) |  |
| **Glucose, mmol/L** |  |  |  |  | <0.001 |  |  | <0.001 |
| <3.9 | 43/323(13.3) | 14/137(10.2) | 29/143(20.3) | 0/21(0) | 0.007 | 2/59(3.4) | 41/242(16.9) | 0.006 |
| 3.9-6.1 | 150/323(46.4) | 85/137(62) | 62/143(43.4) | 3/21(14.3) | <0.001 | 19/59(32.2) | 131/242(54.1) | 0.004 |
| >6.1 | 108/323(33.4) | 38/137(27.7) | 52/143(36.4) | 18/21(85.7) | <0.001 | 38/59(64.4) | 70/242(28.9) | <0.001 |
| **Potassium, mmol/L** |  |  |  |  | 0.022 |  |  | 0.581 |
| <3.5 | 73/323(22.6) | 27/143(18.9) | 42/137(30.7) | 4/21(19) | 0.063 | 15/58(25.9) | 58/243(23.9) | 0.882 |
| 3.5-5.5 | 225/323(69.7) | 116/143(81.1) | 93/137(67.9) | 16/21(76.2) | 0.037 | 42/58(72.4) | 183/243(75.3) | 0.774 |
| >5.5 | 3/323(0.9) | 0/143(0) | 2/137(1.5) | 1/21(4.8) | 0.057 | 1/58(1.7) | 2/243(0.8) | 0.475 |
| **RT-PCR** |  |  |  |  | <0.001 |  |  | <0.001 |
| Positive | 186/323(57.6) | 69/151(45.7) | 95/146(65.1) | 22/26(84.6) | <0.001 | 50/63(79.4) | 136/260(52.3) |  |
| Negative | 137/323(42.4) | 82/151(54.3) | 51/146(34.9) | 4/26(15.4) | <0.001 | 13/63(20.6) | 124/260(47.7) |  |

\*Data were missing for PH in 262(81.1%), for lactate in 266(81.8%), for PaO2 in 262(81.1%), for PaCO2 in 265(82.5%), for PaO2:FiO2 in 264(81.7%)
†Data were missing for creatine kinase in 224(69.3%) for lactate dehydrogenase in 236(73.1%)

**Supplementary Table S2.** Univariate analysis associated with clinical outcome of patients with COVID-19.

|  |  |  |  |  |  |
| --- | --- | --- | --- | --- | --- |
| Variables |  | Univariate Analysis |  |  |  |
|  |  | Odds Ratio | 95%CI |  | p |
| Age | ≥65 | 15.274 | 2.016 | 115.741 | 0.001 |
| Smoking | Yes | 2.118 | 1.002 | 4.474 | 0.049 |
| BMI | ≥30 | 3.604 | 1.154 | 11.256 | 0.009 |
| Hypnotics | Yes | 0.199 | 0.076 | 0.519 | <0.001 |
| Dyspnea | Yes | 3.316 | 1.107 | 9.93 | 0.032 |
| Diabetes | Yes | 3.578 | 1.839 | 6.963 | <0.001 |
| Cardiovascular and cerebrovascular diseases | Yes | 2.154 | 1.043 | 4.449 | 0.038 |
| Malignancy | Yes | 6.45 | 1.054 | 39.456 | 0.044 |
| Serum amyloid A, mg/liter | >10 | 9.356 | 1.253 | 69.852 | 0.029 |
| Procalcitonin, ng/mL | ≥0.1 | 0.252 | 0.074 | 0.852 | 0.027 |
| Hypersensitive troponin I, pg/mL | >0.04 | 3.562 | 1.028 | 12.338 | 0.045 |
| Creatine kinase CMB, U/L | ≥5 | 4.43 | 1.481 | 13.247 | 0.008 |
| Lactate dehydrogenase, U/L | >250 | 5.885 | 1.082 | 32.013 | 0.003 |
| Alanine aminotransferase, U/L |  |  |  |  |  |
|  | <7 | 0.313 | 0.092 | 1.061 | 0.014 |
|  | >40 | 2.083 | 1.09 | 3.979 | 0.002 |
| Aspartate aminotransferase, U/L |  |  |  |  |  |
|  | <13 | 0.525 | 0.174 | 1.584 | 0.024 |
|  | >35 | 3.239 | 1.776 | 5.906 | <0.001 |
| Blood urea nitrogen, mmol/L | >8 | 2.08 | 1.112 | 3.892 | 0.006 |
| Creatinine, μmol/L | <88 | 0.447 | 0.21 | 0.95 | 0.003 |
| Glucose, mmol/L |  |  |  |  |  |
|  | <3.9 | 0.336 | 0.075 | 1.505 | 0.018 |
|  | >6.1 | 3.743 | 2.008 | 6.975 | <0.001 |
| White blood cell count, $\times 10^9/L$ | >10 | 11.871 | 4.51 | 31.245 | <0.001 |
| Neutrophil count, $\times 10^9/L$ | >75 | 9.43 | 4.427 | 20.084 | <0.001 |
| Platelet count, $\times 10^9/L$ | $\geq 150$ | 2.981 | 1.056 | 8.415 | 0.008 |
| PCR at Diagnosis | positive | 2.862 | 1.472 | 5.565 | 0.00194 |
| Disease severity on admission |  |  |  |  |  |
|  | Severe | 1.162 | 0.605 | 2.234 | <0.001 |
|  | Critical | 27.51 | 9.315 | 81.241 | <0.001 |
| Bilateral GGO | Yes | 0.511 | 0.291 | 0.899 | 0.020 |
| Crazy paving sign | Yes | 2.697 | 1.169 | 6.22 | 0.020 |
| Diffuse patchy ground glass and air bronchogram | Yes | 2.542 | 1.067 | 6.056 | 0.035 |
| Multiple Bilateral pulmonary multiple consideration<br>and intralobular interstitial thickening | Yes | 16.112 | 3.261 | 79.602 | <0.001 |

## Notes

### Competing Interest Statement

The authors have declared no competing interest.

